# Association between remnant cholesterol-to-HDL-C ratio and obstructive sleep apnea risk in U.S. adults: a cross-sectional analysis of NHANES 2015–2018

**DOI:** 10.64898/2026.05.09.26352793

**Authors:** Zhiyuan Zhu, Shan Shan

**Author notes:** Correspondence: Zhiyuan Zhu, Graduate School, Hebei Medical University, Shijiazhuang 050017, Hebei, P. R. China, Shan Shan, Bethune International Peace Hospital, Shijiazhuang 050082, Hebei, P. R. China.

## Abstract

**Background:** Several lipid ratios have been linked to obstructive sleep apnea (OSA) risk in NHANES, yet two questions central to clinical translation remain unanswered: how much of the association is carried by central adiposity, and whether the dose–response curve contains an actionable threshold. We addressed both for the remnant cholesterol-to-HDL-C ratio (RC/HDL-C).

**Methods:** We analysed 3,635 adults aged ≥20 years from NHANES 2015–2018. OSA risk was ascertained from the Sleep Disorders Questionnaire. Multivariable logistic regression estimated odds ratios across three nested models. Restricted cubic splines and segmented regression characterised the dose–response and located the inflection point. Mediation by body roundness index (BRI) was quantified by nonparametric percentile bootstrap (1,000 resamples). Discrimination was compared by ROC analysis, with stratified and trimmed-sample sensitivity analyses.

**Results:** OSA risk was identified in 1,361 participants (37.4%). Each one-unit rise in RC/HDL-C carried 23% higher adjusted odds of OSA (OR 1.23, 95% CI 1.03–1.47); the highest quartile carried 49% higher odds than the lowest (P-trend < 0.001). The dose–response was nonlinear, with an inflection at RC/HDL-C = 0.232: below this point each 0.1-unit increase raised odds by 54% (OR 1.54, 95% CI 1.16–2.05); above it the curve plateaued. BRI mediated 82.7% of the total effect (ACME 0.039, P < 0.001), with the indirect pathway 2.8 times stronger in women. AUCs were 0.599 (BRI) and 0.564 (RC/HDL-C).

**Conclusions:** RC/HDL-C showed a modest, threshold-shaped association with OSA risk in U.S. adults, with central adiposity (BRI) as the predominant mediating factor. These exploratory findings, based on questionnaire-defined OSA, warrant prospective validation in cohorts with polysomnography.

## Introduction

Obstructive sleep apnea (OSA) is a common sleep-related breathing disorder characterised by recurrent partial or complete collapse of the upper airway during sleep. Current estimates place the global burden at close to one billion adults aged 30–69 years, of whom roughly 425 million have moderate-to-severe disease [1]. The clinical consequences extend well beyond daytime sleepiness: intermittent hypoxia, fragmented sleep and heightened sympathetic drive together promote oxidative stress, low-grade inflammation and metabolic disturbance, and OSA maintains bidirectional relationships with hypertension, type 2 diabetes, cardiovascular disease and the metabolic syndrome [2–4]. Undiagnosed and untreated OSA erodes quality of life, reduces work capacity and imposes substantial economic costs[5], so identifying people at high risk early — and with inexpensive tools — remains a pressing clinical need.

Lipid metabolism is one of the pathways through which OSA exerts its systemic effects[6]. Intermittent hypoxia activates the sympathetic nervous system and inflammatory cascades, suppresses lipoprotein lipase activity, and slows the clearance of triglyceride-rich lipoproteins, allowing remnant particles to accumulate [7]. Remnant cholesterol (RC) — defined as the cholesterol carried by atherogenic lipoproteins other than LDL and HDL — is closely tied to low-grade inflammation and endothelial dysfunction [8]. HDL cholesterol (HDL-C), in contrast, has anti-inflammatory, antioxidant and endothelial-protective properties, and low concentrations are a familiar marker of dyslipidaemia [9]. Because a single lipid species captures only one side of this biology, ratios that combine a proatherogenic with a protective component have gained traction in recent cardiometabolic work.

Among these, the RC/HDL-C ratio is attractive for two reasons. It incorporates the proinflammatory burden of remnant lipoproteins and the protective role of HDL-C in a single number, and it is computed from variables that appear on almost every routine lipid panel. The ratio has been linked to type 2 diabetes, the metabolic syndrome and incident cardiovascular events [10,11], but whether it tracks OSA risk in a large, nationally representative sample has not been addressed.

Obesity is a well-established driver of OSA: fat deposition around the neck and in the pharyngeal walls narrows the upper airway, while abdominal and thoracic fat restrict diaphragmatic excursion and reduce lung volume [12]. Body mass index (BMI) is still the default measure in clinical practice, yet BMI does not discriminate between central and peripheral fat distribution. The body roundness index (BRI) is derived from waist circumference and height and more faithfully reflects visceral adiposity and overall body shape [13]. Given the physiological links between lipid dysregulation and fat accumulation, BRI is a plausible mediator on any pathway from RC/HDL-C to OSA — but this hypothesis has not, to our knowledge, been formally tested.

Using cross-sectional data from the 2015–2018 cycles of the National Health and Nutrition Examination Survey (NHANES), we set out to quantify the association between RC/HDL-C and OSA risk, to describe its dose–response shape and any threshold behaviour, and to ask how much of the association is carried by central adiposity as measured by BRI. Our aim was to provide evidence that might support the use of inexpensive, widely available markers for OSA risk stratification.

## Materials and methods

### Study design and data source

This was a cross-sectional analysis of data from NHANES, a continuous programme run by the National Center for Health Statistics (NCHS) that uses a complex, multistage, stratified probability design to collect demographic, health, dietary and laboratory information from a nationally representative sample of the U.S. population. The NHANES protocol was approved by the NCHS Research Ethics Review Board and all participants gave written informed consent. Because the present analysis uses publicly available, de-identified secondary data, no further ethical approval was required. We combined the 2015–2016 and 2017–2018 cycles because these were the two most recent cycles that contained the sleep questionnaire, lipid panel and anthropometric measurements required for the analysis.

### Study population

A total of 19,225 participants were enrolled in NHANES 2015–2018. We applied four exclusion criteria in sequence: (1) age below 20 years (n = 7,937); (2) missing lipid measurements (total cholesterol, LDL-C or HDL-C), waist circumference or height (n = 6,913); (3) incomplete sleep-questionnaire data (n = 26); and (4) missing values for any key covariate (n = 714). The final analytic sample comprised 3,635 adults (Figure 1).

**Figure 1.**
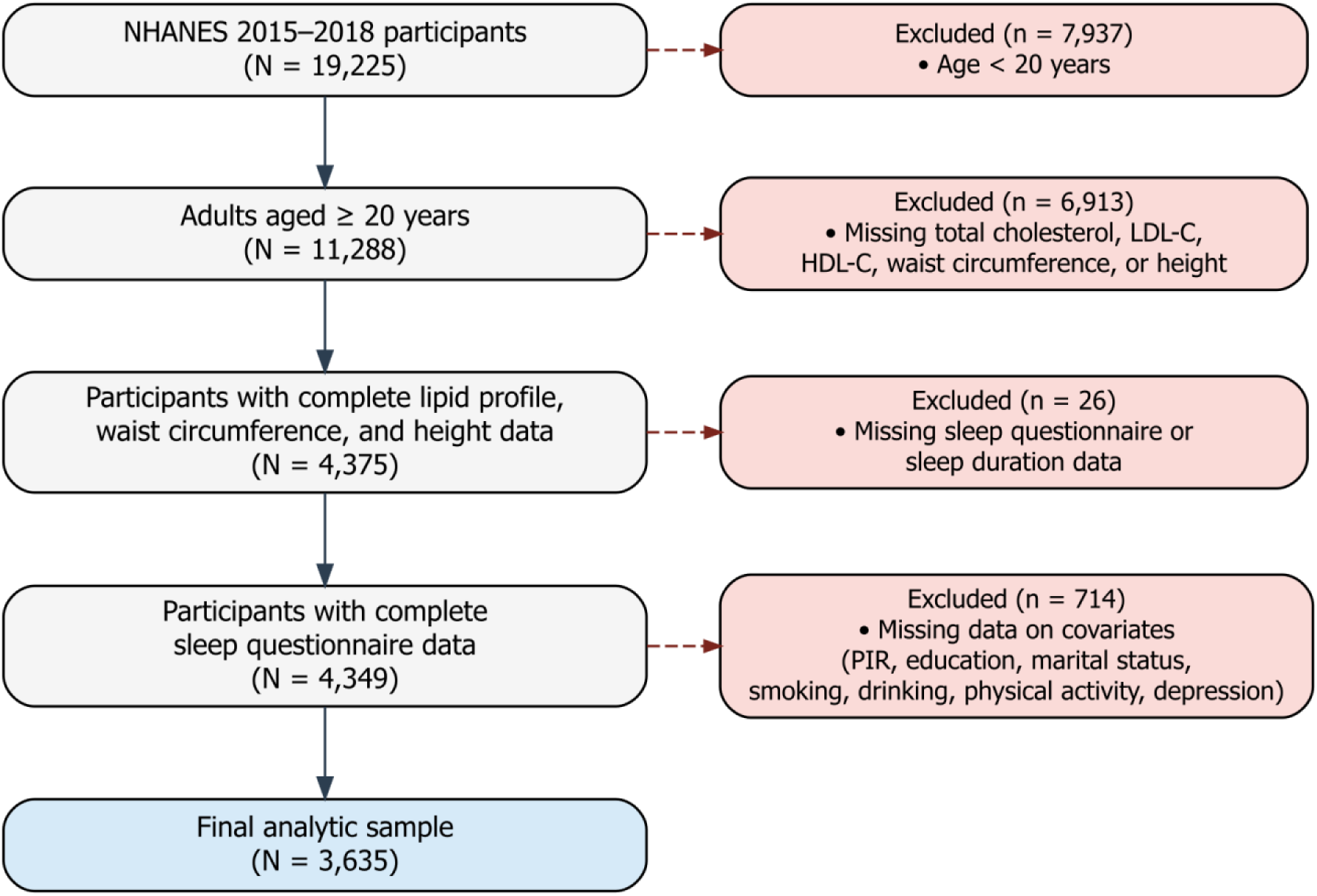
Flow diagram of participant selection from NHANES 2015–2018.

### Exposure

RC/HDL-C ratio

Lipid testing was carried out at NHANES-contracted laboratories using standardised assays. Total cholesterol (TC), LDL-C and HDL-C are reported in mg/dL.

Remnant cholesterol was computed as:

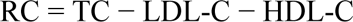

The exposure was then defined as the ratio RC/HDL-C.

#### Mediator

body roundness index (BRI)

BRI was derived from waist circumference (WC, in metres) and height (H, in metres) using the formula proposed by Thomas and colleagues [11]:

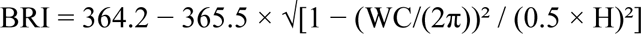

BRI captures central adiposity and visceral fat distribution more accurately than BMI.

### Outcome

#### OSA risk

Because NHANES does not conduct polysomnography (PSG), we classified OSA risk from the Sleep Disorders Questionnaire, consistent with earlier NHANES-based epidemiological studies [14]. Participants were considered at risk if they reported habitual snoring (SLQ030) or witnessed apnea (SLQ040) on at least three nights per week, or excessive daytime sleepiness (SLQ120) on 16–30 days per month, in conjunction with a self-reported weekday sleep duration of at least seven hours. Because this classification is questionnaire-based rather than derived from the PSG gold standard, we consistently refer to the outcome as “OSA risk” throughout.

#### Covariates

Informed by prior NHANES analyses, ten covariates were included: age (continuous), sex (male/female), race/ethnicity (Mexican American, Other Hispanic, Non-Hispanic White, Non-Hispanic Black, Other), education (five categories), marital status (six categories), poverty-to-income ratio (PIR, continuous), smoking status (never, former, current), alcohol use (yes/no, based on ALQ130), physical activity (yes/no, derived from PAQ650 and PAQ665), and depression (PHQ-9 total ≥ 10)[15].

BMI, hypertension, diabetes mellitus and cardiovascular disease were deliberately not adjusted for. BMI is strongly collinear with BRI, and adjusting for both would obscure the physiological meaning of the mediator. The three chronic conditions plausibly lie downstream of the RC/HDL-C → OSA pathway; controlling for downstream variables would amount to overadjustment and bias the mediation estimates.

#### Statistical analysis

All analyses were performed in R 4.5.3, drawing mainly on the mediation, rms, pROC and segmented packages. A two-sided P < 0.05 was treated as statistically significant. Continuous variables are summarised as mean ± standard deviation or median [interquartile range] according to their distribution; categorical variables are given as counts and percentages. Participants were grouped into quartiles of RC/HDL-C, and between-group differences were tested with the chi-square test or the Kruskal–Wallis test as appropriate.

Three nested logistic regression models were fitted. Model 1 was unadjusted. Model 2 adjusted for age, sex, race/ethnicity, education, marital status and PIR. Model 3 additionally adjusted for smoking, alcohol use, physical activity and depression. RC/HDL-C was analysed both as a continuous variable and in quartiles, and a linear trend across quartiles was tested.

Mediation was assessed within the Baron–Kenny framework [16] using a nonparametric percentile bootstrap with 1,000 resamples implemented in the mediation R package [17]; we report the average causal mediation effect (ACME), the average direct effect (ADE), the total effect and the proportion mediated, each with 95% confidence intervals. Primary mediation analyses used the fully adjusted model, and additional analyses were stratified by sex and age.

The dose–response shape was examined with restricted cubic splines (RCS) with knots placed at the 5th, 35th, 65th and 95th percentiles[18]. When nonlinearity was detected, segmented regression was used to locate the inflection point and to estimate the slopes on either side.

Discrimination of RC/HDL-C and BRI for OSA risk was compared using ROC curves; differences in the area under the curve (AUC) were tested with the DeLong method, and optimal cut-offs were selected by the Youden index[19].

Subgroup analyses were prespecified for age group (20–39, 40–59, ≥ 60 years), sex, race/ethnicity, BRI tertile, smoking, alcohol use and depression, with interaction tested by likelihood-ratio tests. To evaluate robustness to extreme values, all mediation analyses were repeated after excluding participants with RC/HDL-C values in the top and bottom 1%.

## Results

### Baseline characteristics

The analytic sample comprised 3,635 adults with a mean age of 50.3 ± 17.4 years; 1,784 (49.1%) were men and 1,851 (50.9%) were women. Participants were grouped into quartiles of RC/HDL-C: Q1 (n = 910), Q2 (n = 908), Q3 (n = 915) and Q4 (n = 902). Overall, 1,361 (37.4%) screened positive for OSA risk. Baseline characteristics are shown in Table 1.

**Table 1.** Basic Characteristics of Research Subjects.

|  | Characteristics | Overall | Q1 | Q2 | Q3 | Q4 | P value |
| --- | --- | --- | --- | --- | --- | --- | --- |
| n |  | 3635 | 910 | 908 | 915 | 902 |  |
| Age,years(mean (SD)) |  | 50.31 (17.35) | 47.09 (18.36) | 50.41 (17.82) | 52.60 (16.92) | 51.12 (15.72) | <0.001 |
| Age group,n(%) | 20-39 | 1140 (31.4) | 380 (41.8) | 281 (30.9) | 230 (25.1) | 249 (27.6) | <0.001 |
|  | 40-59 | 1215 (33.4) | 260 (28.6) | 289 (31.8) | 321 (35.1) | 345 (38.2) |  |
|  | ≥60 | 1280 (35.2) | 270 (29.7) | 338 (37.2) | 364 (39.8) | 308 (34.1) |  |
| Sex,n(%) | Male | 1784 (49.1) | 339 (37.3) | 400 (44.1) | 491 (53.7) | 554 (61.4) | <0.001 |
|  | Female | 1851 (50.9) | 571 (62.7) | 508 (55.9) | 424 (46.3) | 348 (38.6) |  |
| Race,n(%) | Mexican American | 547 (15.0) | 89 (9.8) | 132 (14.5) | 151 (16.5) | 175 (19.4) | <0.001 |
|  | Other Hispanic | 412 (11.3) | 71 (7.8) | 97 (10.7) | 119 (13.0) | 125 (13.9) |  |
|  | Non-Hispanic White | 1328 (36.5) | 308 (33.8) | 303 (33.4) | 349 (38.1) | 368 (40.8) |  |
|  | Non-Hispanic Black | 754 (20.7) | 281 (30.9) | 242 (26.7) | 149 (16.3) | 82 (9.1) |  |
|  | Other Race - Including Multi-Racial | 594 (16.3) | 161 (17.7) | 134 (14.8) | 147 (16.1) | 152 (16.9) |  |
| Education level,n(%) | Less than 9th grade | 297 (8.2) | 54 (5.9) | 71 (7.8) | 79 (8.6) | 93 (10.3) | <0.001 |
|  | 9-11th grade (Includes 12th grade with no diploma) | 415 (11.4) | 77 (8.5) | 101 (11.1) | 126 (13.8) | 111 (12.3) |  |
|  | High school graduate/GED or equivalent | 848 (23.3) | 196 (21.5) | 208 (22.9) | 222 (24.3) | 222 (24.6) |  |
|  | Some college or AA degree | 1159 (31.9) | 292 (32.1) | 307 (33.8) | 276 (30.2) | 284 (31.5) |  |
|  | College graduate or above | 916 (25.2) | 291 (32.0) | 221 (24.3) | 212 (23.2) | 192 (21.3) |  |
| Marital status,n(%) | Married | 1862 (51.2) | 411 (45.2) | 448 (49.3) | 492 (53.8) | 511 (56.7) | <0.001 |
|  | Widowed | 241 (6.6) | 60 (6.6) | 66 (7.3) | 67 (7.3) | 48 (5.3) |  |
|  | Divorced | 434 (11.9) | 93 (10.2) | 115 (12.7) | 103 (11.3) | 123 (13.6) |  |
|  | Separated | 121 (3.3) | 27 (3.0) | 29 (3.2) | 30 (3.3) | 35 (3.9) |  |
|  | Never married | 637 (17.5) | 229 (25.2) | 164 (18.1) | 127 (13.9) | 117 (13.0) |  |
|  | Living with partner | 340 (9.4) | 90 (9.9) | 86 (9.5) | 96 (10.5) | 68 (7.5) |  |
| PIR (median [IQR]) |  | 2.12 [1.18, 4.02] | 2.39 [1.24, 4.40] | 2.08 [1.22, 3.86] | 2.11 [1.11, 3.87] | 2.06 [1.15, 3.70] | 0.004 |
| PIR,n(%) | Low | 1043 (28.7) | 237 (26.0) | 248 (27.3) | 286 (31.3) | 272 (30.2) | 0.004 |
|  | Medium | 1514 (41.7) | 358 (39.3) | 392 (43.2) | 375 (41.0) | 389 (43.1) |  |
|  | High | 1078 (29.7) | 315 (34.6) | 268 (29.5) | 254 (27.8) | 241 (26.7) |  |
| Smoking status,n(%) | never | 2014 (55.4) | 564 (62.0) | 536 (59.0) | 486 (53.1) | 428 (47.5) | <0.001 |
|  | former | 927 (25.5) | 203 (22.3) | 213 (23.5) | 243 (26.6) | 268 (29.7) |  |
|  | current | 694 (19.1) | 143 (15.7) | 159 (17.5) | 186 (20.3) | 206 (22.8) |  |
| Alcohol drinking,n(%) | No | 1111 (30.6) | 249 (27.4) | 278 (30.6) | 288 (31.5) | 296 (32.8) | 0.075 |
|  | Yes | 2524 (69.4) | 661 (72.6) | 630 (69.4) | 627 (68.5) | 606 (67.2) |  |
| Physical activity,n(%) | Inactive | 1878 (51.7) | 396 (43.5) | 445 (49.0) | 500 (54.6) | 537 (59.5) | <0.001 |
|  | Active | 1757 (48.3) | 514 (56.5) | 463 (51.0) | 415 (45.4) | 365 (40.5) |  |
| Depression,n(%) | No | 3325 (91.5) | 842 (92.5) | 840 (92.5) | 845 (92.3) | 798 (88.5) | 0.003 |
|  | Yes | 310 (8.5) | 68 (7.5) | 68 (7.5) | 70 (7.7) | 104 (11.5) |  |
| BRI (median [IQR]) |  | 5.30[3.99, 7.03] | 4.11 [2.99, 5.57] | 5.19 [3.86, 6.65] | 5.78 [4.60, 7.54] | 6.11 [4.85, 7.82] | <0.001 |
| BMI (median [IQR]) |  | 28.40 [24.60, 33.50] | 25.20 [22.00, 29.50] | 27.90 [24.00, 32.35] | 29.50 [26.10, 34.60] | 30.80 [27.10, 35.30] | <0.001 |
| OSA risk,n(%) | No OSA risk | 2274 (62.6) | 641 (70.4) | 570 (62.8) | 548 (59.9) | 515 (57.1) | <0.001 |
|  | OSA risk | 1361 (37.4) | 269 (29.6) | 338 (37.2) | 367 (40.1) | 387 (42.9) |  |

Continuous variables are expressed as mean ± standard deviation or median [IQR]; categorical variables are expressed as frequency (%). p-values were calculated by chi-square test or Kruskal-Wallis test.

Several characteristics varied systematically across quartiles. Mean age rose from 47.1 ± 18.4 years in Q1 to 52.6 ± 16.9 years in Q3 (P < 0.001), and the proportion of men increased from 37.3% in Q1 to 61.4% in Q4 (P < 0.001). The racial distribution also shifted: the share of Mexican Americans rose from 9.8% to 19.4% across quartiles, while that of Non-Hispanic Black participants fell from 30.9% to 9.1% (P < 0.001).

College graduates became less common at higher quartiles (32.0% in Q1 vs 21.3% in Q4; P < 0.001), whereas the married proportion was highest in Q4 (56.7%) and the never-married proportion lowest (13.0%; P < 0.001). Low-income participants were over-represented in Q4 (30.2%; P = 0.004). Lifestyle profiles grew less favourable with higher RC/HDL-C: current smoking rose from 15.7% to 22.8% and former smoking from 22.3% to 29.7% (P < 0.001); physical inactivity increased from 43.5% to 59.5% (P < 0.001); alcohol use did not differ significantly between groups (P = 0.075); and depression was most common in Q4 (11.5%; P = 0.003).

Anthropometric indices tracked RC/HDL-C closely. Median BRI rose from 4.11 in Q1 to 6.11 in Q4 (P < 0.001), and median BMI from 25.2 to 30.8 kg/m² (P < 0.001). The prevalence of positive OSA risk increased monotonically from 29.6% in Q1 to 42.9% in Q4 (P < 0.001), pointing to a positive association between RC/HDL-C and OSA risk that warranted formal testing.

### Association between RC/HDL-C and OSA risk

Logistic regression (Table 2) showed an independent, dose-responsive positive association between RC/HDL-C and OSA risk. In the unadjusted model, each one-unit increase in RC/HDL-C was associated with a 45% rise in the odds of OSA (OR 1.45, 95% CI 1.22–1.72; P < 0.001). Adjusting for demographic factors attenuated the estimate to OR 1.27 (95% CI 1.06–1.52), and after further adjustment for lifestyle and psychological factors in Model 3 the association persisted (OR 1.23, 95% CI 1.03–1.47).

**Table 2.**
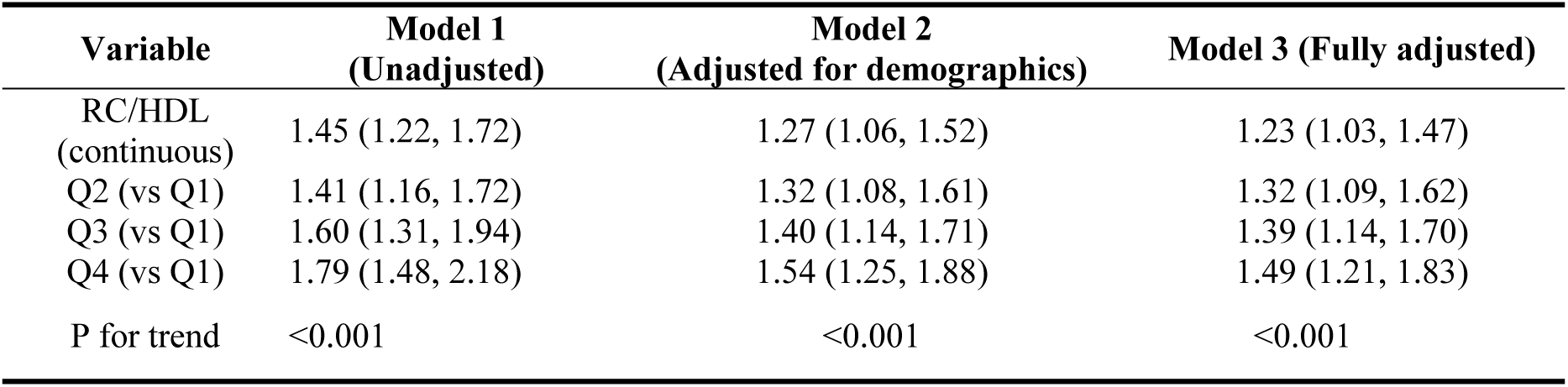
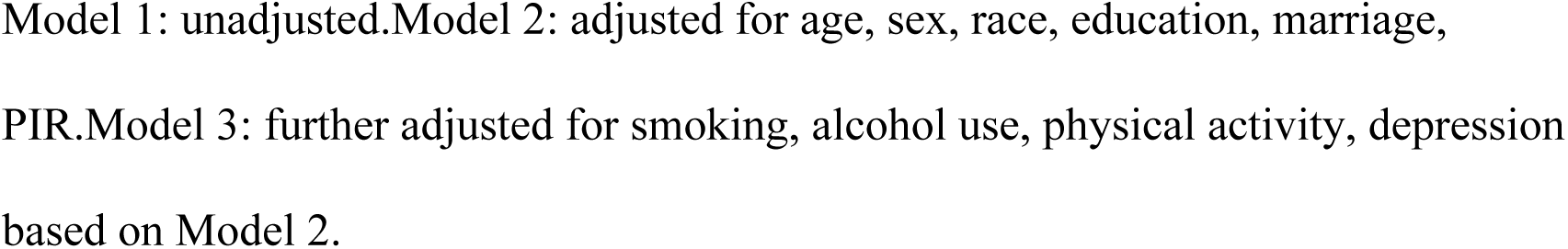
Logistic Regression Analysis of RC/HDL and OSA Risk Association.

| Variable | Model 1<br>(Unadjusted) | Model 2<br>(Adjusted for demographics) | Model 3 (Fully adjusted) |
| --- | --- | --- | --- |
| RC/HDL<br>(continuous) | 1.45 (1.22, 1.72) | 1.27 (1.06, 1.52) | 1.23 (1.03, 1.47) |
| Q2 (vs Q1) | 1.41 (1.16, 1.72) | 1.32 (1.08, 1.61) | 1.32 (1.09, 1.62) |
| Q3 (vs Q1) | 1.60 (1.31, 1.94) | 1.40 (1.14, 1.71) | 1.39 (1.14, 1.70) |
| Q4 (vs Q1) | 1.79 (1.48, 2.18) | 1.54 (1.25, 1.88) | 1.49 (1.21, 1.83) |
| P for trend | <0.001 | <0.001 | <0.001 |

Model 1: unadjusted. Model 2: adjusted for age, sex, race, education, marriage, PIR. Model 3: further adjusted for smoking, alcohol use, physical activity, depression based on Model 2.

Quartile analyses gave consistent results. Compared with Q1, the adjusted odds ratios in Q2, Q3 and Q4 were 1.32 (95% CI 1.09–1.62), 1.39 (95% CI 1.14–1.70) and 1.49 (95% CI 1.21–1.83), respectively; participants in the highest quartile thus had 49% higher odds of OSA than those in the lowest. The trend across quartiles was highly significant in all three models (P for trend < 0.001), supporting a stable dose–response relationship.

### Nonlinear dose–response and threshold effect

After full adjustment, RCS analysis revealed a clearly nonlinear relationship between RC/HDL-C and OSA risk (P-overall < 0.001; P-nonlinear < 0.001; Figure 2). The curve rose steeply at low values and levelled off at higher values. Segmented regression placed the inflection at RC/HDL-C = 0.232 (95% CI 0.170–0.294). Below this point, each 0.1-unit increase in RC/HDL-C was associated with 54% higher odds of OSA (OR 1.54, 95% CI 1.16–2.05; P = 0.003); above it, further increases had no appreciable effect (OR 1.00, 95% CI 0.98–1.02; P = 0.770).

**Figure 2.**
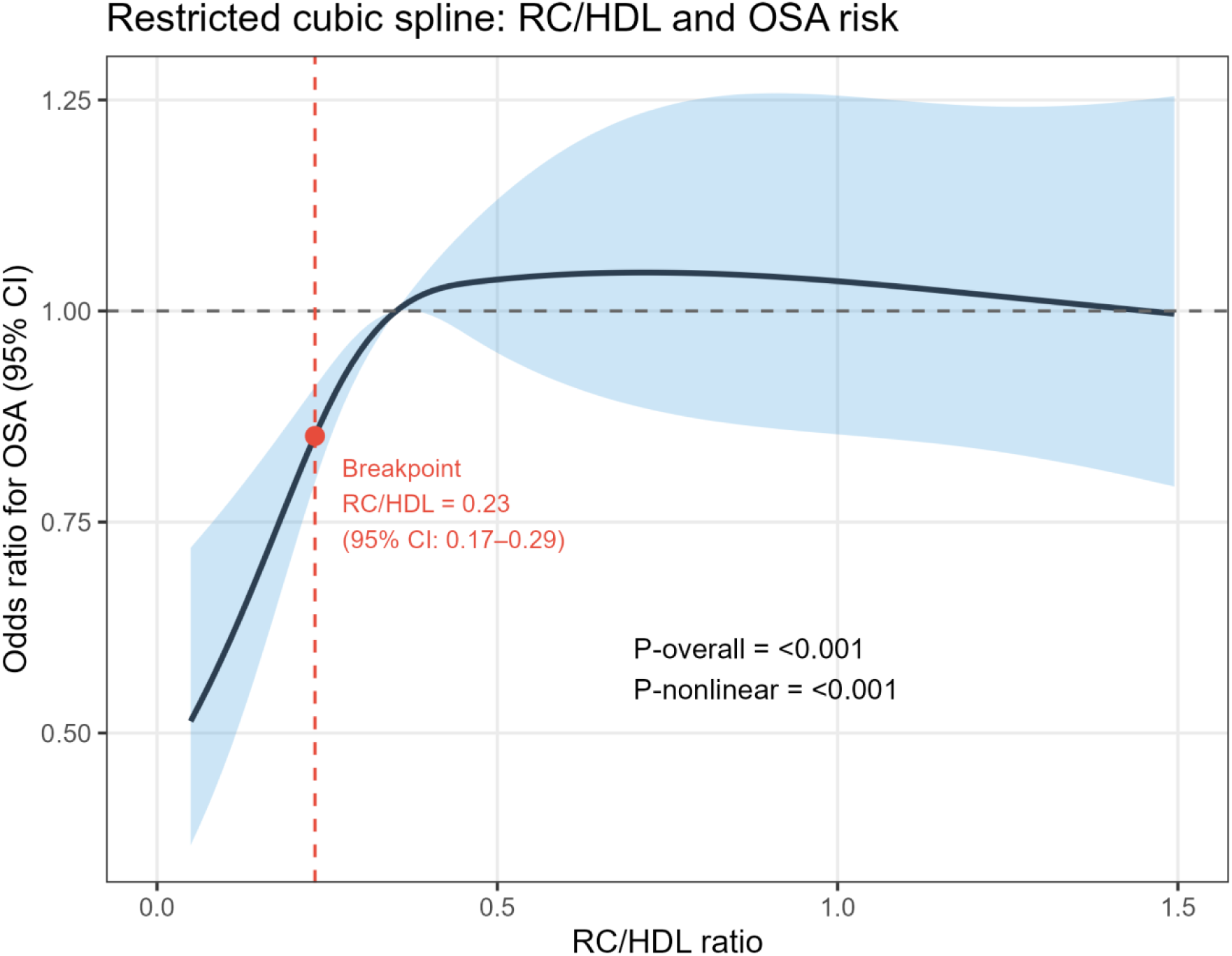
Restricted cubic spline (RCS) analysis of the association between RC/HDL-C and OSA risk. The solid line shows the adjusted odds ratio and the shaded area the 95% confidence interval (Model 3: adjusted for age, sex, race/ethnicity, education, marital status, PIR, smoking, alcohol use, physical activity and depression). The red dashed vertical line marks the inflection point at RC/HDL-C = 0.23.

Two readings of this threshold pattern are plausible. One is physiological compensation: above a certain level of metabolic disturbance, additional lipid dysregulation does not translate into further increments in OSA risk. The other is that once RC/HDL-C is high, OSA risk comes to be dominated by factors that are downstream of — or independent from — the lipid ratio itself, including central adiposity and systemic inflammation. Practically, the finding suggests that RC/HDL-C values close to the inflection point may carry more prognostic information than very high values, which already sit on the plateau.

### BRI mediates the RC/HDL-C–OSA association

To probe the mechanism, we fitted a mediation model with BRI as the mediator, adjusting for age, sex, race/ethnicity, education, marital status, PIR, smoking, alcohol use, physical activity and depression. Results are shown in Table 3 and Figure 3. Both legs of the indirect pathway were significant. A one-unit increase in RC/HDL-C was associated with a 1.383-unit increase in BRI (path a: β = 1.383, P < 0.001), and higher BRI was in turn associated with greater OSA risk (path b: β = 0.124, P < 0.001). The total effect of RC/HDL-C on OSA risk was significant (total effect 0.047, 95% CI 0.006–0.099; P = 0.026), and was driven almost entirely by the indirect path through BRI (ACME 0.039, 95% CI 0.028–0.052; P < 0.001). The direct effect was small and not significant (ADE 0.008, 95% CI −0.032 to 0.059; P = 0.700). BRI therefore accounted for 82.7% of the total effect (95% CI 36.4–336.5%; P = 0.026), indicating that most of the impact of RC/HDL-C on OSA risk is transmitted through central adiposity.

**Figure 3.**
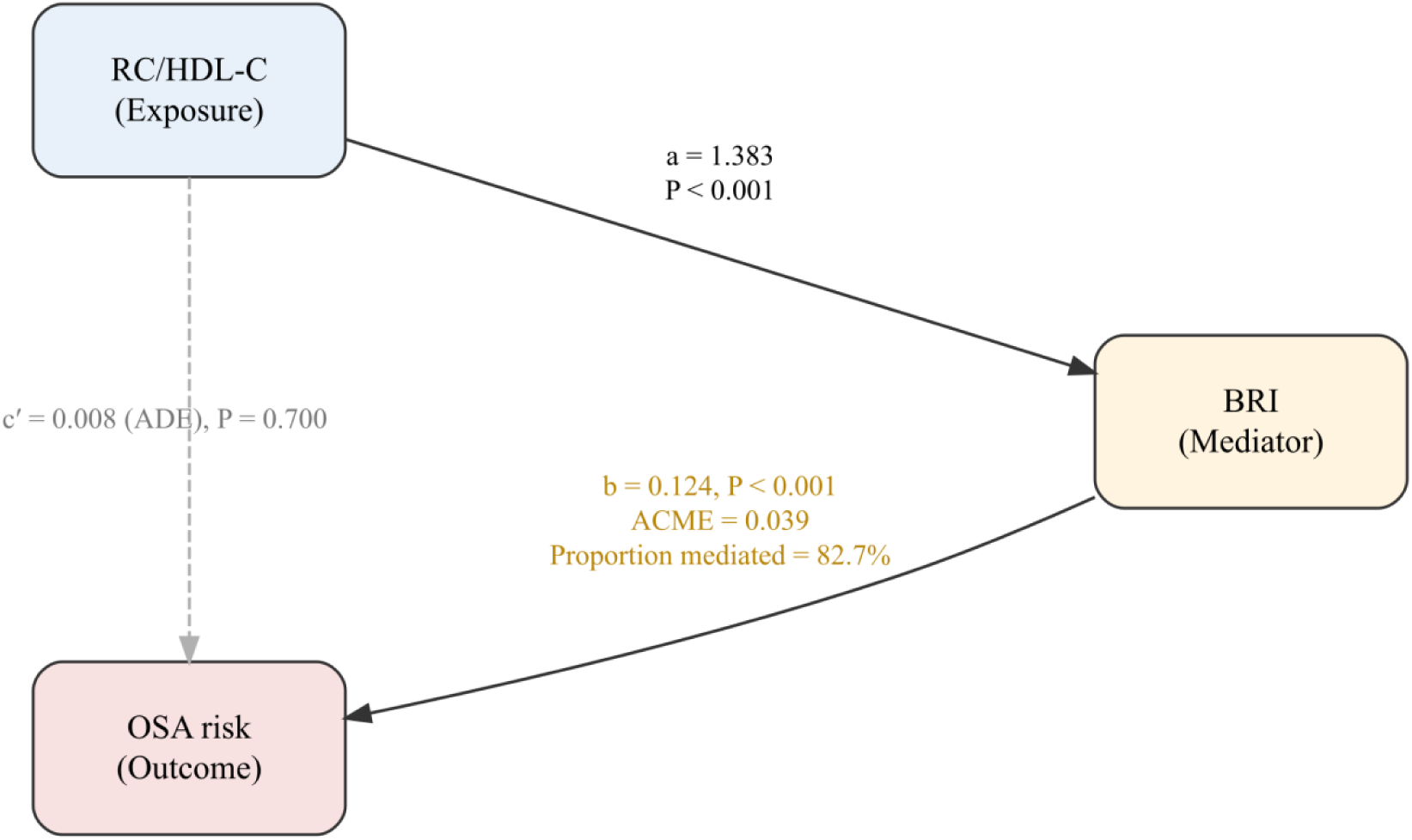
Path diagram of the mediation model. RC/HDL-C is the exposure, OSA risk the outcome and BRI the mediator. Path a: effect of RC/HDL-C on BRI; path b: effect of BRI on OSA risk; path c: total effect of RC/HDL-C on OSA risk; path c′: direct effect of RC/HDL-C on OSA risk not mediated by BRI.

**Table 3.** BRI mediation analysis of the association between RC/HDL and OSA risk.

| Effect | Estimate (log-odds) | 95% CI | P value |
| --- | --- | --- | --- |
| ACME (indirect) | 0.039 | (0.027, 0.052) | <0.001 |
| ADE (direct) | 0.008 | (-0.032, 0.059) | 0.700 |
| Total effect | 0.047 | (0.006, 0.099) | 0.026 |
| Proportion mediated | 82.7% | (36.4%, 336.5%) | 0.026 |

Models adjusted for age, sex, race, education, marriage, PIR, smoking, alcohol consumption, physical activity, depression. acme: average causal mediated effect; ade: average direct effect. 95% CI estimated by nonparametric percentile Bootstrap method (1000 replications).

### Subgroup mediation

To check whether the mediation effect was consistent across strata, we repeated the analysis within sex and age subgroups (Table 4). Mediation through BRI was significant in both men and women, but the indirect effect was markedly larger in women (ACME 0.064, 95% CI 0.045–0.087; proportion mediated 68.3%) than in men (ACME 0.023, 95% CI 0.011–0.038; proportion mediated 106.4%). The women-to-men ratio of about 2.8 suggests that the RC/HDL-C → BRI → OSA pathway is more pronounced in women, which is biologically plausible given sex differences in fat distribution and in the regulation of lipid metabolism by sex hormones.

**Table 4.** .Subgroup mediation analysis results.

| Subgroup | N | ACME (95% CI) | Mediation Proportion, %<br>(95% CI) | ACME P value |
| --- | --- | --- | --- | --- |
| Male | 1784 | 0.023 (0.011, 0.038) | 106.4 (-982.8, 1072.5) | <0.001 |
| Female | 1851 | 0.064 (0.045, 0.087) | 68.3 (37.2, 227.8) | <0.001 |
| Younger (<60 years old) | 2355 | 0.041 (0.028, 0.059) | 90.9 (-305.5, 659.3) | <0.001 |
| Older (≥60 years old) | 1280 | 0.037 (0.018, 0.060) | 107.8(-764.3, 1012.6) | <0.001 |

Stratification variables have been removed from the model (gender is not adjusted when stratified by sex; age is not adjusted when stratified by age). acme: average causal mediated effect; mp: mediated proportion.

Across age strata, the indirect effect was similar: ACME 0.041 (95% CI 0.028–0.059) in those under 60 years and 0.037 (95% CI 0.018–0.060) in those aged 60 or older. The corresponding proportions mediated were 90.9% and 107.8%. Confidence intervals for the proportion mediated were wide in some strata, reflecting a well-known mathematical instability that arises when the direct effect is close to zero; the ACME estimates, however, were stable across all strata and confirm the robustness of the mediation.

### Subgroup analyses and interaction tests

Subgroup analyses across seven prespecified effect modifiers are summarised in Figure 4 and Table 5. The direction of association was positive in most strata, although statistical significance varied with subgroup size. The association was strongest in adults aged 20–39 (OR 1.41, 95% CI 1.00–2.00; P = 0.050), in women (OR 1.51, 95% CI 1.10–2.07; P = 0.010), and in Non-Hispanic Black participants (OR 1.78, 95% CI 1.06–3.05; P = 0.030). It remained significant among alcohol users (OR 1.24, 95% CI 1.01–1.53; P = 0.047) and among participants without depression (OR 1.26, 95% CI 1.04–1.53; P = 0.018). Stratifying by BRI tertile dissolved the association within each tertile (all P > 0.05), which is consistent with the mediation findings: once participants are matched on central adiposity, little residual effect of RC/HDL-C on OSA risk remains. None of the seven interaction tests was significant (age P = 0.301; sex P = 0.109; race P = 0.220; BRI P = 0.400; smoking P = 0.427; alcohol P = 0.726; depression P = 0.310), indicating that the overall effect of RC/HDL-C on OSA risk did not differ meaningfully across these subgroups.

**Figure 4.**
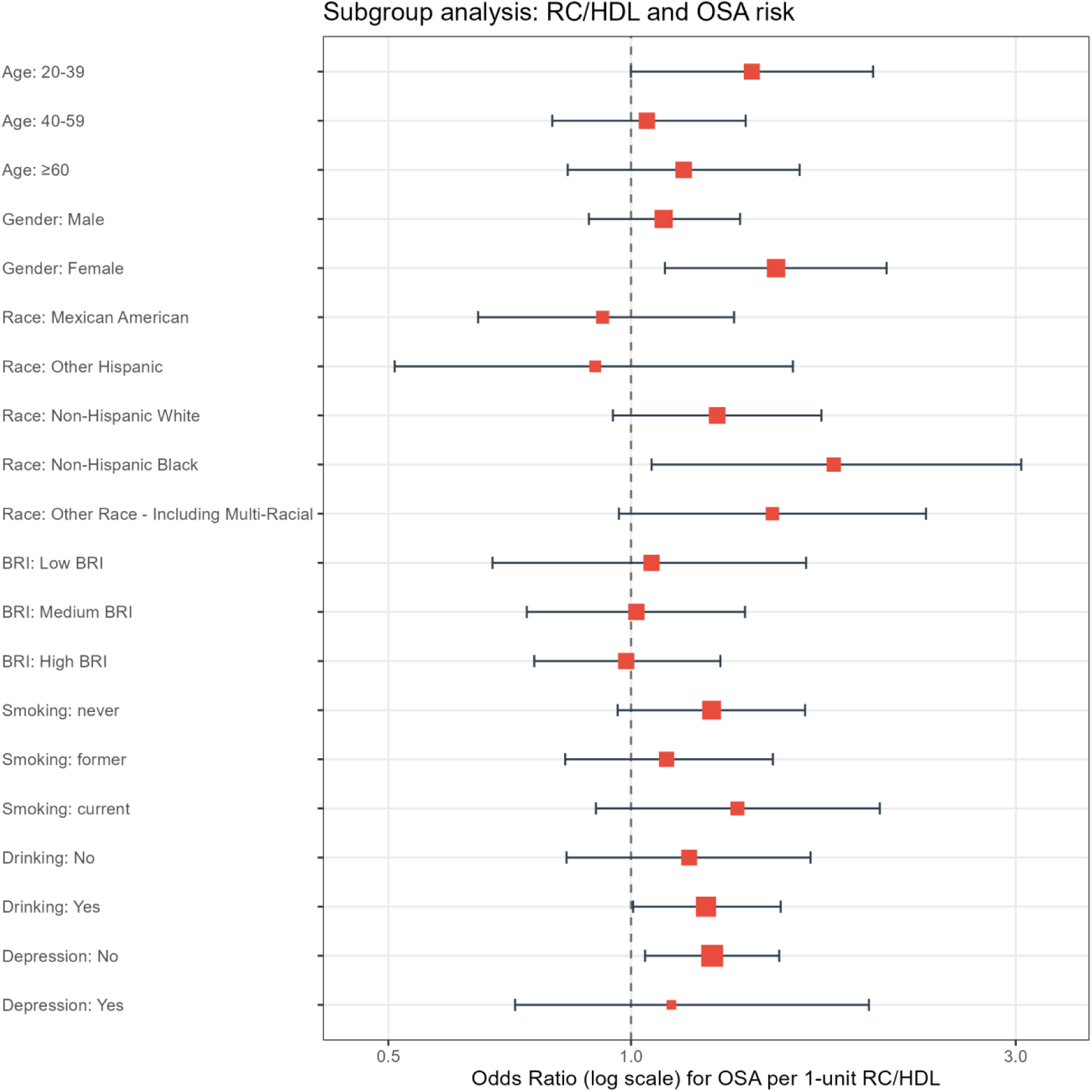
Forest plot of subgroup analyses for the association between RC/HDL-C and OSA risk.

**Table 5.**
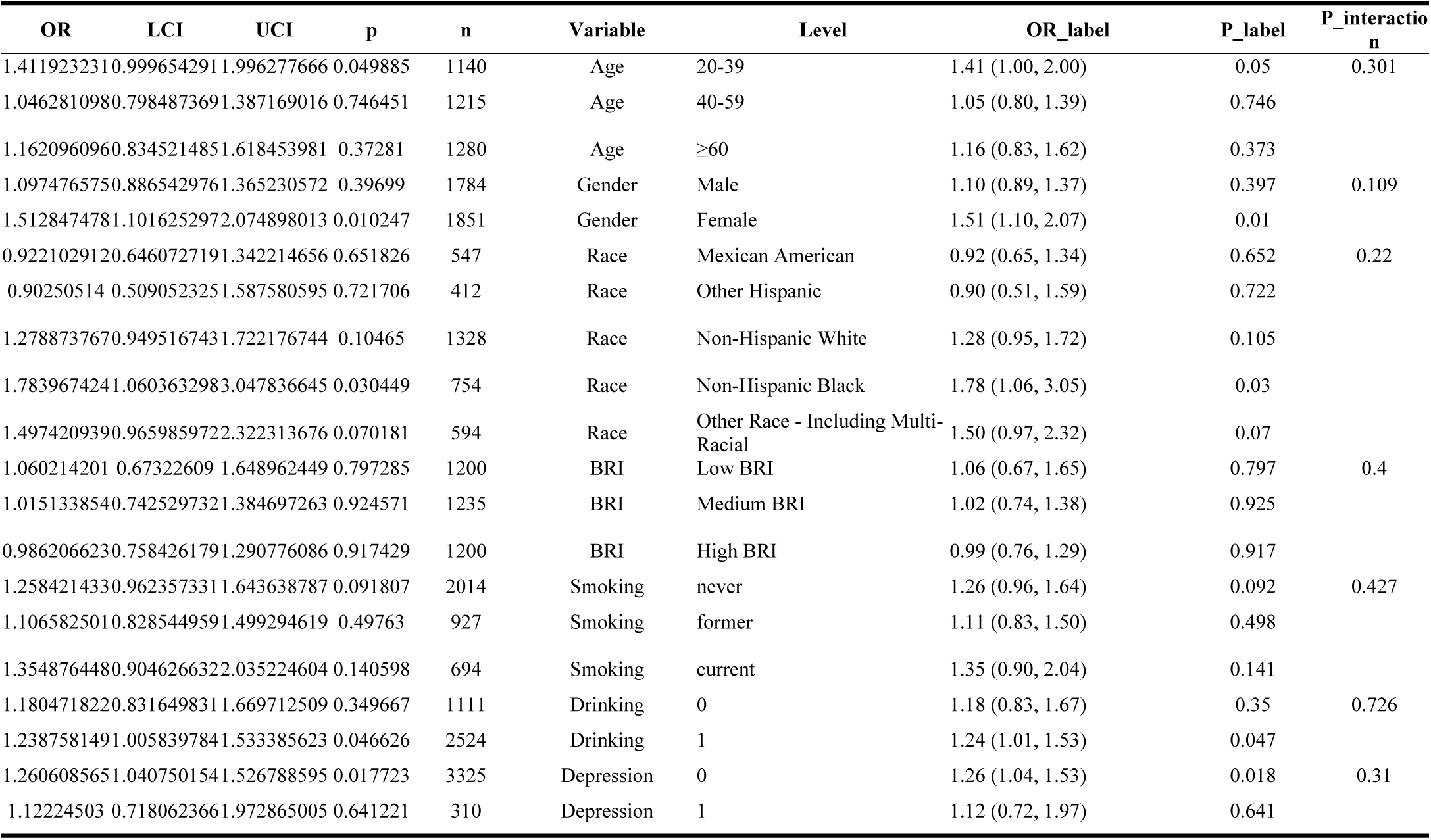
Subgroup analysis of RC/HDL and OSA risk.

### Discrimination: RC/HDL-C versus BRI

ROC analysis (Figure 5, Table 6) compared the discriminative performance of the two indices. BRI showed slightly better discrimination than RC/HDL-C: AUC 0.599 (95% CI 0.580–0.618) for BRI, with an optimal cut-off of 0.29 (sensitivity 63.2%, specificity 52.2%, Youden index 0.154), versus AUC 0.564 (95% CI 0.545–0.583) for RC/HDL-C, with an optimal cut-off of 0.326 (sensitivity 60.2%, specificity 49.7%, Youden index 0.099). Neither 95% CI crossed 0.5, so both indices offer some, though only modest, discriminative power. The AUC advantage of about 0.035 in favour of BRI is consistent with the mediation estimate that BRI carries the bulk of the association, and reinforces the interpretation of BRI as the principal mediator on the pathway from RC/HDL-C to OSA.

**Figure 5.**
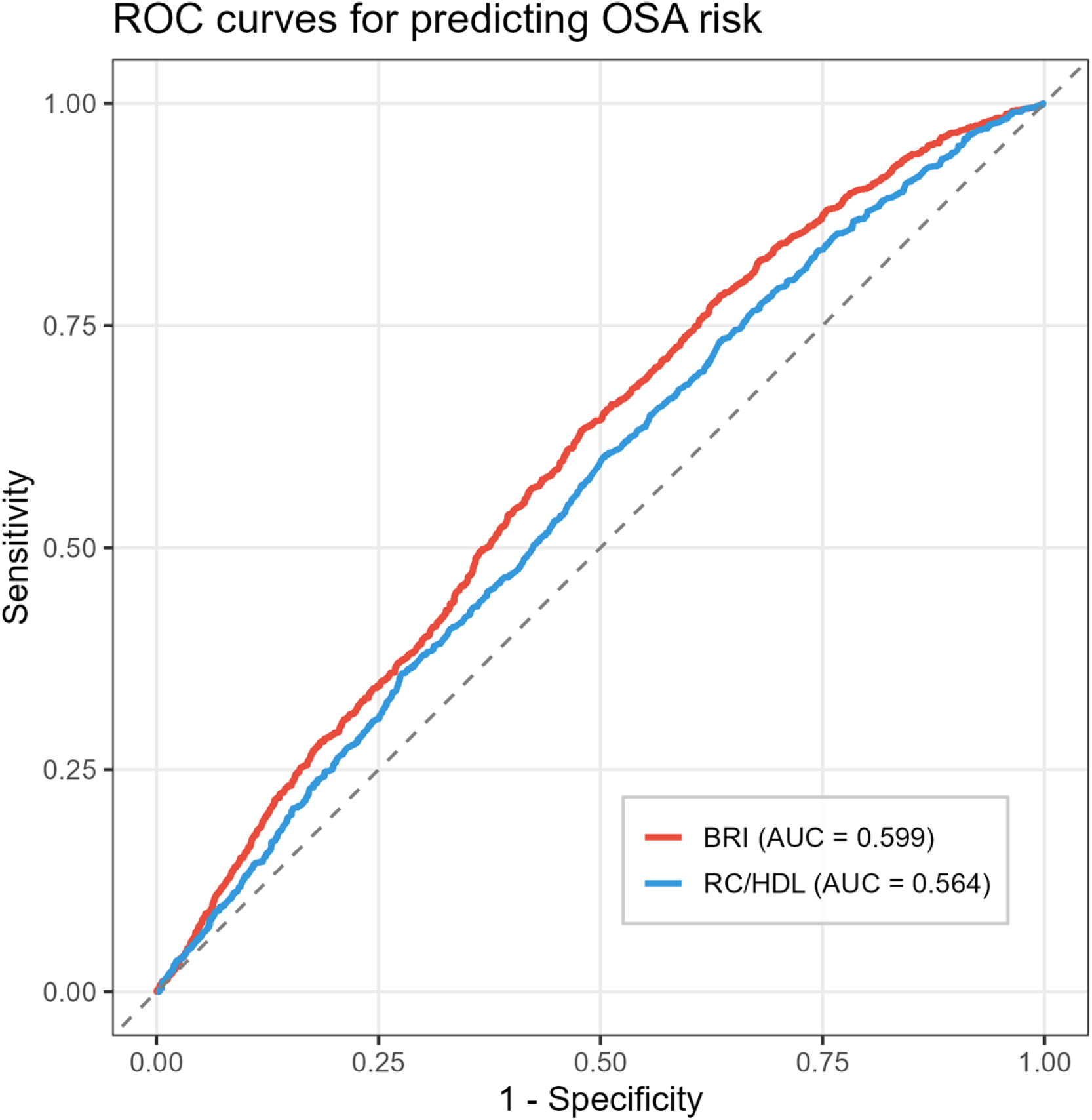
ROC curves of RC/HDL-C and BRI for predicting OSA risk.

**Table 6.** ROC curve analysis results.

| Marker | AUC | 95% CI | Optimal Cutoff | Sensitivity | Specificity | Youden Index |
| --- | --- | --- | --- | --- | --- | --- |
| RC/HDL | 0.564 | (0.545, 0.583) | 0.326 | 0.602 | 0.497 | 0.099 |
| BRI | 0.599 | (0.580, 0.618) | 0.29 | 0.632 | 0.522 | 0.154 |

The optimal cutoff value was determined based on the Youden index. p-value for the difference in AUC was calculated by the DeLong test.

### Sensitivity analysis

To test whether the main results were driven by extreme lipid values, we excluded participants with RC/HDL-C values in the top or bottom 1% (n = 72), leaving 3,563 adults. After trimming (Table 7), the total effect was somewhat larger than in the primary analysis (total effect 0.074, 95% CI 0.030–0.122; P = 0.006), and the BRI-mediated indirect effect remained robust (ACME 0.049, 95% CI 0.036–0.064; P < 0.001). The proportion mediated was 66.9% (95% CI 36.6–165.2%; P = 0.006), with a noticeably narrower confidence interval than in the primary analysis. The direct effect was again non-significant (ADE 0.024, 95% CI −0.022 to 0.074; P = 0.334). The direction of every estimate matched the primary analysis, indicating that the conclusion — that RC/HDL-C influences OSA risk largely through BRI — is not sensitive to extreme values.

**Table 7.** Sensitivity analysis: mediation analysis results after excluding extreme RC/HDL values.

| Indicator | Main analysis (N=3635) | Sensitivity analysis (N=3563) |
| --- | --- | --- |
| Total effect | 0.047 (0.006, 0.099), p=0.026 | 0.074 (0.030, 0.122), p=0.006 |
| ACME (indirect) | 0.039 (0.027, 0.052), p=<0.001 | 0.049(0.036, 0.064), p=<0.001 |
| ADE (direct) | 0.008 (-0.032, 0.059), p=0.700 | 0.024 (-0.022, 0.074), p=0.334 |
| Proportion mediated | 82.7% (36.4%, 336.5%), p=0.026 | 66.9% (36.6%, 165.2%), p=0.006 |

Sensitivity analyses excluded participants with RC/HDL outside the 1st and 99th percentiles (N = 3,563).

## Discussion

Using a nationally representative sample of U.S. adults, we examined the link between the RC/HDL-C ratio and OSA risk and mapped the mechanistic route by which this association operates. Three findings stand out. First, RC/HDL-C is independently associated with OSA risk, with roughly half again the odds of OSA in the highest quartile relative to the lowest. Second, the dose–response is nonlinear, with an inflection near RC/HDL-C = 0.23 beyond which the association plateaus. Third, central adiposity indexed by BRI carries the bulk of the association — about 82.7% in the primary analysis — while the direct effect of the lipid ratio is small and non-significant. Subgroup and sensitivity analyses support the robustness of these findings.

### Possible mechanisms linking RC/HDL-C to OSA

An elevated RC/HDL-C ratio reflects two concurrent shifts — a rise in remnant lipoproteins and a fall in HDL-C — which together mark generalised dyslipidaemia. Remnant lipoproteins are rich in triglycerides and cholesterol, are taken up by monocytes and macrophages, accumulate in vascular walls and activate inflammatory pathways including NF-κB and IL-6 [20]. In the upper airway, chronic inflammation contributes to mucosal oedema, reduced muscle tone and local fat infiltration, all of which predispose to sleep-related airway collapse [21]. HDL-C normally counteracts these processes through its anti-inflammatory, antioxidant and endothelial-protective actions, but the intermittent hypoxia that characterises OSA favours oxidative modification of HDL particles and compromises their function [22]. A ratio that summarises both the proinflammatory and the protective facets of the lipid profile therefore captures aspects of OSA-relevant metabolism that a single marker cannot.This convergence of inflammation, oxidative stress and lipid remodelling provides a coherent mechanistic backdrop linking RC/HDL-C to OSA pathogenesis [23,24].

### A biological reading of the threshold

The inflection at RC/HDL-C ≈ 0.23 is worth dwelling on. Below this point, a 0.1-unit increment was associated with roughly 54% higher odds of OSA, so the system seems highly sensitive to small perturbations when lipid derangement is mild. Above the threshold, risk plateaus. One plausible reading is compensation: sustained elevation of RC/HDL-C may upregulate antioxidant defences or recruit alternative pathways that partially offset further lipid-related damage [25]. Another is that, once metabolic disturbance is severe, OSA risk comes to be dominated by mechanical drivers — neck-fat accumulation and airway geometry — rather than by the lipid ratio itself. From a screening perspective, the threshold suggests that individuals whose RC/HDL-C sits just above the inflection may be the most informative subgroup to monitor, because they are past the steepest part of the risk curve but have not yet reached the plateau.

### Central adiposity as the key mediator

That BRI accounted for about 82.7% of the total effect while the direct effect was negligible underscores the central place of body-fat distribution in the pathway from dyslipidaemia to OSA. Disordered lipid metabolism promotes visceral and neck fat accumulation, which in turn increases waist circumference, compresses upper-airway soft tissue and limits diaphragmatic excursion, all of which lower functional lung volumes and predispose the airway to collapse during sleep [26,27]. We chose BRI rather than BMI as the mediator because BRI, by combining waist circumference with height, captures central obesity and overall body roundness more directly than BMI; prior work also found that BRI outperforms BMI in its association with OSA [28].Recent work using other waist-based indices (e.g., visceral adiposity index, weight-adjusted waist index) has shown comparable patterns in NHANES populations [29].

### Subgroup differences

The indirect effect through BRI was about 2.8 times larger in women than in men. This is consistent with the known role of oestrogen in steering fat to peripheral depots (gluteofemoral) and in maintaining higher HDL-C concentrations; an elevated RC/HDL-C in women often signals that this protective pattern has given way, with a parallel shift to central fat deposition [30]. The pathway from RC/HDL-C through BRI to OSA therefore becomes more conspicuous in women. Reassuringly, none of the seven interaction tests reached significance, so the overall direction and magnitude of the RC/HDL-C–OSA association were broadly similar across strata, supporting the generalisability of the main findings.These observations align with broader epidemiological evidence that OSA in women is more strongly tied to central adiposity and post-menopausal metabolic shifts than in men [31].

### Discrimination and clinical implications

BRI discriminated OSA risk slightly better than RC/HDL-C (AUC 0.599 vs 0.564), but neither index approached the conventional threshold for useful screening (AUC > 0.7). This is in line with earlier cross-sectional studies of lipid ratios in OSA [32], and reinforces the point that no single biochemical or anthropometric measure will replace a comprehensive clinical evaluation. Combining BRI and RC/HDL-C, or integrating them into established screening instruments such as the STOP-BANG questionnaire, may be a more realistic strategy than relying on either marker alone[33].

### Strengths and limitations

This study has several strengths. To the best of our knowledge, it is the first to examine the RC/HDL-C–OSA relationship in a nationally representative adult population, and it combines logistic regression, RCS-based nonlinearity testing, segmented regression, formal mediation analysis, ROC comparison, subgroup and interaction testing, and a sensitivity analysis. The convergence of these approaches supports the robustness of the findings.

Several limitations should nevertheless be acknowledged. First, the cross-sectional design precludes causal inference. OSA itself, through intermittent hypoxia, can alter lipid metabolism, so reverse causation cannot be ruled out; longitudinal cohorts and Mendelian randomisation will be needed to clarify the direction of effect[34]. Second, OSA risk was ascertained from a self-reported questionnaire rather than PSG, the diagnostic gold standard, which introduces a degree of misclassification even though this approach has been used in prior NHANES-based work[35]. Third, NHANES lacks detailed information on dietary fat intake, lipid-lowering medication use and genetic variants, all of which can influence RC/HDL-C. Fourth, BMI, hypertension, diabetes and cardiovascular disease were not included as covariates for the reasons set out in the Methods; while we consider this choice methodologically defensible, we did not perform a formal sensitivity analysis to quantify the impact of alternative adjustment strategies, and this remains a trade-off inherent to our mediation framework.

### Conclusion

In the NHANES 2015–2018 adult population, the RC/HDL-C ratio was independently and positively associated with OSA risk, with a clear threshold effect at RC/HDL-C ≈ 0.23. Most of this association — about 83% — was mediated by central adiposity as measured by BRI, identifying body-fat distribution as the principal bridge between the lipid ratio and OSA. Because RC/HDL-C is computed from variables that appear on any standard lipid panel, combining it with a simple body-shape index may offer a practical, low-cost way to flag adults at elevated OSA risk and to guide interventions aimed at improving lipid metabolism and reducing central obesity. Prospective studies and mechanistic work are now needed to test whether RC/HDL-C and BRI, used together, improve the performance of existing OSA screening tools.

## Data Availability

No data were generated by this study. All data analysed are publicly available from the National Health and Nutrition Examination Survey (NHANES) at https://wwwn.cdc.gov/nchs/nhanes/. Derived datasets and analytic code are available from the corresponding author on reasonable request.

## Abbreviations

ACME: Average causal mediation effect
ADE: Average direct effect
AUC: Area under the curve
BMI: Body mass index
BRI: Body roundness index
CI: Confidence interval
CVD: Cardiovascular disease
HDL-C: High-density lipoprotein cholesterol
LDL-C: Low-density lipoprotein cholesterol
NCHS: National Center for Health Statistics
NHANES: National Health and Nutrition Examination Survey
OR: Odds ratio
OSA: Obstructive sleep apnea
PHQ-9: Patient Health Questionnaire-9
PIR: Poverty-to-income ratio
PSG: Polysomnography
RC: Remnant cholesterol
RCS: Restricted cubic spline
ROC: Receiver operating characteristic
TC: Total cholesterol.

## Acknowledgements

We thank all NHANES participants and the National Center for Health Statistics for making these data publicly available.

## Authors’ contributions

Zhiyuan Zhu conceived and designed the study, curated the data, performed the formal analysis and interpretation, drafted the manuscript and revised it critically for important intellectual content. Shan Shan supervised the study, validated the analyses, and revised the manuscript critically for important intellectual content. Both authors approved the final version for publication and agree to be accountable for all aspects of the work.

## Funding

This research received no specific grant from any funding agency in the public, commercial or not-for-profit sectors.

## Availability of data and materials

The data analysed here are publicly available from NHANES at https://wwwn.cdc.gov/nchs/nhanes/. Derived datasets used in this study are available from the corresponding author on reasonable request.

## Declarations

### Ethics approval and consent to participate

This study is a secondary analysis of publicly available, de-identified data from NHANES 2015--2018. The research was conducted in accordance with the Declaration of Helsinki. The NHANES protocol was approved by the NCHS Research Ethics Review Board, and all original participants provided written informed consent. Additional ethical approval for the present retrospective analysis was waived because the data are publicly accessible and fully anonymised.

### Consent for publication

Not applicable.

### Consent to participate

Not applicable.

### Competing interests

The authors declare that they have no competing interests.

## Notes

### Competing Interest Statement

The authors have declared no competing interest.

### Funding Statement

The author(s) received no specific funding for this work.

### Author Declarations

This study is a secondary analysis of publicly available, de-identified data from the National Health and Nutrition Examination Survey (NHANES) 2015–2018. The original NHANES protocol was approved by the National Center for Health Statistics (NCHS) Research Ethics Review Board, and all participants provided written informed consent. Additional ethical approval for the present retrospective analysis was waived because the data are publicly accessible and fully anonymised.

